# A multi-omics resource of B cell activation reveals genetic mechanisms for immune-mediated diseases

**DOI:** 10.1101/2025.05.22.25328104

**Authors:** Vitor R. C. Aguiar, Marcella E. Franco, Nada Abdel Aziz, Daniela Fernandez-Salinas, Marcos Chiñas, Mariasilvia Colantuoni, Qian Xiao, Nicolaj Hackert, Yifei Liao, Rodrigo Cervantes-Diaz, Marc Todd, Brian Wauford, Alex Wactor, Vaishali Prahalad, Raquel Laza-Briviesca, Roxane Darbousset, Qiang Wang, Scott Jenks, Kevin S. Cashman, Esther Zumaquero, Zhu Zhu, Junning Case, Paloma Cejas, Miguel Gomez, Hannah Ainsworth, Miranda Marion, Mehdi Benamar, Pui Lee, Lauren Henderson, Margaret Chang, Kevin Wei, Henry Long, Carl D. Langefeld, Benjamin E. Gewurz, Ignacio Sanz, Jeffrey A. Sparks, Esra Meidan, Peter A. Nigrovic, Maria Gutierrez-Arcelus

## Abstract

Most genetic variants that confer risk of complex immune-mediated diseases (IMDs) affect gene regulation in specific cell types. Their target genes and focus cell types are often unknown, partially because some effects are hidden in untested cell states. B cells play central roles in IMDs, including autoimmune, allergic, infectious, and cancer-related diseases. Despite this established importance, B cell activation states are underrepresented in functional genomics studies. In this study, we obtained B cells from 26 healthy female donors and stimulated them *in vitro* with six activation conditions targeting key pathways: the B cell receptor (BCR), Toll-like receptor 7 (TLR7), TLR9, CD40, and a cocktail that promotes differentiation into double negative 2 (DN2) IgD^-^ CD27^-^ CD11c^+^ CD21^-^ B cells, a likely pathogenic subset implicated in autoimmunity and infection. We profiled up to 24 B cell activation states and up to 5 control conditions using RNA-seq, single-cell RNA-seq with surface protein markers (CITE-seq), and ATAC-seq. We characterize how IMD-associated genes respond to stimuli and group into distinct functional programs. High-depth RNA-seq data reveals widespread splicing effects during B cell activation. Using single-cell data, we describe stimulus-dependent B cell fates. Chromatin data reveal transcription factors likely involved in B cell activation, and activation-dependent open chromatin regions that are enriched in IMD genetic risk. We experimentally validate a lupus risk variant in a stimulus-specific open chromatin region that regulates *TNFSF4* expression, highlighting the relevance of studying B cell activation to elucidate disease association. These data are shared via an interactive browser that can be used to query the dynamics of gene regulation and B cell differentiation during activation by different stimuli, enhancing further investigation of B cells and their role in IMDs: https://mgalab.shinyapps.io/bcellactivation.

## Introduction

B lymphocytes are essential players in the immune system, with their ability to produce antibodies, present antigens, and release cytokines. While these functions are critical for protecting the host from infections and maintaining immune homeostasis, dysregulated B cell activity can drive the development and progression of immune-mediated diseases (IMDs). B cells are highly dynamic, transitioning through various activation and differentiation states in response to diverse external stimuli. Signals from the B cell receptor (BCR), co-stimulatory molecules, cytokines, and pattern recognition receptors (PRRs) converge to define the activation trajectory of B cells. This capacity to reach diverse activation states is critical for immune defense, but when dysregulated, it can also drive autoimmunity or contribute to the development of allergies and cancer.^1–3^ Understanding how B cells transition into pathogenic states is critical for discerning their role in disease progression and developing targeted therapeutic interventions. One key example of a likely pathogenic B cell state is the double negative 2 (DN2) subset, characterized by absence of CD27 and IgD expression and presence of CD11c expression (with some groups also defining them as CD21^-^CD11c^+^ B cells).^4,5^ DN2 B cells are considered a pre-plasmablast state that might emerge through an extra-follicular pathway,^4^ an alternative maturation pathway independent of germinal center dynamics.^6^ These cells are expanded in the peripheral blood of patients with systemic lupus erythematosus (SLE), rheumatoid arthritis (RA), multiple sclerosis (MS), autoimmune anemia, systemic sclerosis (SSc), Sjogren’s disease, Crohn’s disease (CD), and severe SARS-CoV-2 infection.^4^,^7^–^9^ DN2 B cells have also been identified within tumor-infiltrating B cells in over 15 cancer types.^10^ In mice, CD11c^+^ B cells accumulate with age (a.k.a. age-associated B cells, ABCs) and can cause autoimmunity.^9^,^11^,^12^

Understanding how disease-risk genetic variants alter B cell functions to cause complex IMDs is crucial for comprehending disease pathogenesis and developing new therapies. IMDs arise from an imbalance in immune regulation, driven by genetic and environmental factors.^13^ Genome-wide association studies (GWAS) have helped identify hundreds of genetic variants that confer susceptibility to complex diseases. The majority of these variants are located in non-coding regulatory regions of the genome rather than in protein-coding sequences.^14^,^15^ This observation suggests that they exert their effects by modulating gene regulation rather than directly altering protein structure.

Efforts to map these regulatory variants to their gene targets have provided critical insights into the mechanisms underlying IMDs. However, these efforts are often complicated by the cell-type specificity of regulatory elements. For many risk variants, the gene targets and cell types in which they exert their effects remain unknown. This knowledge gap persists partly because most studies have focused on steady-state conditions, leaving specific activation states of immune cells underexplored. It is increasingly recognized that some regulatory elements and risk variants may only become active under particular states of cellular activation.^13^,^16^ A systematic investigation of immune activation states and their associated pathways is therefore essential to uncover these context-specific effects.

Among the immune cell types implicated in IMDs, B cells play a central role in disease pathogenesis. Interestingly, many genetic risk variants associated with IMDs are enriched in regulatory elements that are specific to B cells compared to other immune cell types. For instance, studies have shown that risk variants for SLE, MS, SSc, primary biliary cholangitis (PBC), psoriasis, and CD are highly enriched in acetylated histone regions marking cis-regulatory elements specific to B cells.^17^, ^18^ Similarly, risk variants for SLE, RA, and other IMDs are enriched in open chromatin regions in B cells.^19^ Further, B cell-specific expression quantitative trait loci (eQTLs) are enriched for risk variants associated with SLE, MS, and other IMDs.^20^ These findings highlight the importance of B cell regulatory mechanisms in driving disease susceptibility.

Despite their pivotal role in IMDs, B cells remain underrepresented in functional genomics studies. This is due to their lower frequency in peripheral blood compared to other immune cells, and the technical challenges associated with culturing and studying them *in vitro*. These limitations have hindered efforts to fully understand how distinct B cell states contribute to disease development and progression, and how genetic risk variants operate in B cells and shape their regulatory and transcriptional responses. To date, a comprehensive analysis of how B cells respond dynamically to diverse stimuli over time and transition into pathogenic states is still lacking, presenting a significant gap in our understanding of their role in IMDs.

In this study, we address these challenges by employing a multi-omics approach to dissect chromatin remodeling, gene expression, and B cell differentiation in response to various activation pathways. By targeting key immune signaling pathways, we have systematically profiled B cells in distinct activation states. This multi-omics resource provides a detailed map of the dynamic changes occurring in B cells during activation, offering new opportunities to investigate the transcriptomic and epigenomic “wiring” of B cells and how genetic variants associated with IMDs influence B cell biology.

## Results

### Multi-omics profiling of B cell activation

We recruited 26 unrelated healthy donors from the Mass General Brigham Biobank (MGBB) and the Boston Children’s Hospital Biorepository of Adult Healthy Controls, and enriched for CD19^+^ cells from peripheral blood (Figure S1a). We stimulated these B cells in six activation conditions and two control conditions, and profiled samples at different time points with bulk RNA-seq, bulk ATAC-seq, single-cell RNA-seq, and CITE-seq (Figure 1).

**Figure 1:**
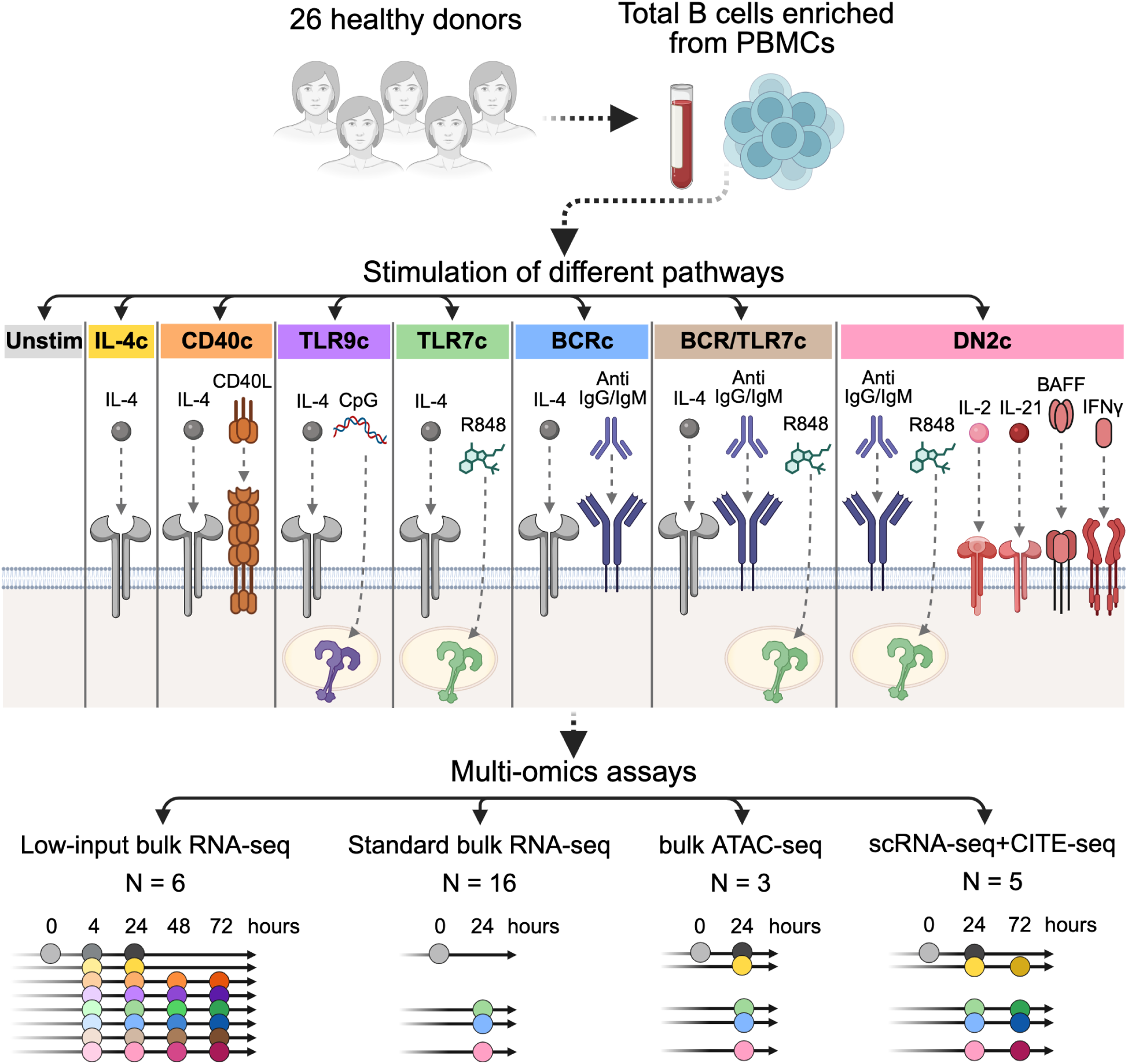
Experimental design and depiction of B cell activation resource. Illustration of experimental design, activation conditions used, and assays performed. At the bottom panel, indicated are the number of donors (N) used for each assay, and the activation or control conditions used, which are color coded as indicated in the middle panel. The three donors for which there is ATAC-seq data were profiled for standard bulk RNA-seq as well. “0 hours” indicates untouched isolated B cells not put on media. Gray conditions of 4 and 24 hours indicate B cells in media only (“Unstim”). Created with BioRender.com.

We used the following stimulation conditions in order to target different activation pathways important in B cells: **(1) CD40c**: CD40 ligand (CD40L) to trigger CD40 signaling, which *in vivo* is done by interaction with T cells and possibly other cell types, along with IL-4 to improve B cell survival;^21^ **(2) TLR9c**: CpG oligodeoxynucleotides to stimulate TLR9, a receptor primarily responsible for detecting bacterial DNA, mimicking bacterial infection,^22^ combined with IL-4;^23^ **(3) TLR7c**: resiquimod (R848) to activate TLR7, which recognizes RNA ligands of viral origin, simulating viral infection, along with IL-4;^23^ **(4) BCRc**: anti-IgG/IgM antibodies to activate the B cell receptor (BCR) signaling pathway, together with IL-4;^19^,^23^ **(5) BCR/TLR7c**: dual BCR and TLR7 stimulation with anti-IgG/IgM, R848, supplemented with IL-4; **(6) DN2c**: a DN2 B cell-inducing cocktail that includes dual BCR and TLR7 stimulation with anti-IgG/IgM and R848, along with additional cytokines (IL-2, IL-21, BAFF, and IFN-γ), which together have been shown to differentiate naive B cells into the “double negative 2” subset (IgD^-^ CD27^-^ CD11c^+^) (Figure S1d,e).^4^ IL-4 was not added to “DN2c”. We verified that each of these activation conditions up-regulated activation marker CD69 with flow cytometry (Figure S1b,c).

Additionally, we used two control conditions: **Unstim** which includes unstimulated B cells at 0 hours as well as cells in media only for 4 and 24 hours (we did not include later time points due to excessive cell death if B cells are not stimulated); and **IL-4c**, which includes media and IL-4 (Figure 1).

### Dynamic gene expression programs in activated B cells

Using low-input RNA-seq, we profiled the *in vitro* activated B cells at multiple time points (0, 4, 24, 48, 72 hours) from 6 adult healthy individuals (Figure 1). Principal component analysis (PCA) on the top 2,000 most variable genes shows that samples group by their activation cell state (Figure 2a). PC1 separates samples to a great extent by the time points, with the 4-hour time point forming one branch, the 24-hour time point forming a second branch, and the 48 and 72-hour time points clustering closer together in a third branch. PC2 strongly separates samples by the *in vitro* conditions with control conditions at the top, followed by CD40c, TLR7c, and TLR9c activation conditions, which are separated from the bottom conditions that include BCR stimulation (BCRc, BCR/TLR7c, DN2c) (Figure 2a, inset). In line with the PCA, pairwise differential expression analysis shows higher numbers of differentially expressed genes for 4 vs. 24 hours than for 48 vs. 72 hours, as well as higher number of differentially expressed genes with respect to control conditions in the three BCR stimulation conditions than in the non-BCR stimulation conditions (Figure 2b). These results suggest that in studies investigating B cell activation response of genes of interest, the time point ascertained matters and that stimulating the BCR itself induces a larger transcriptomic response than stimulating other receptors alone.

**Figure 2:**
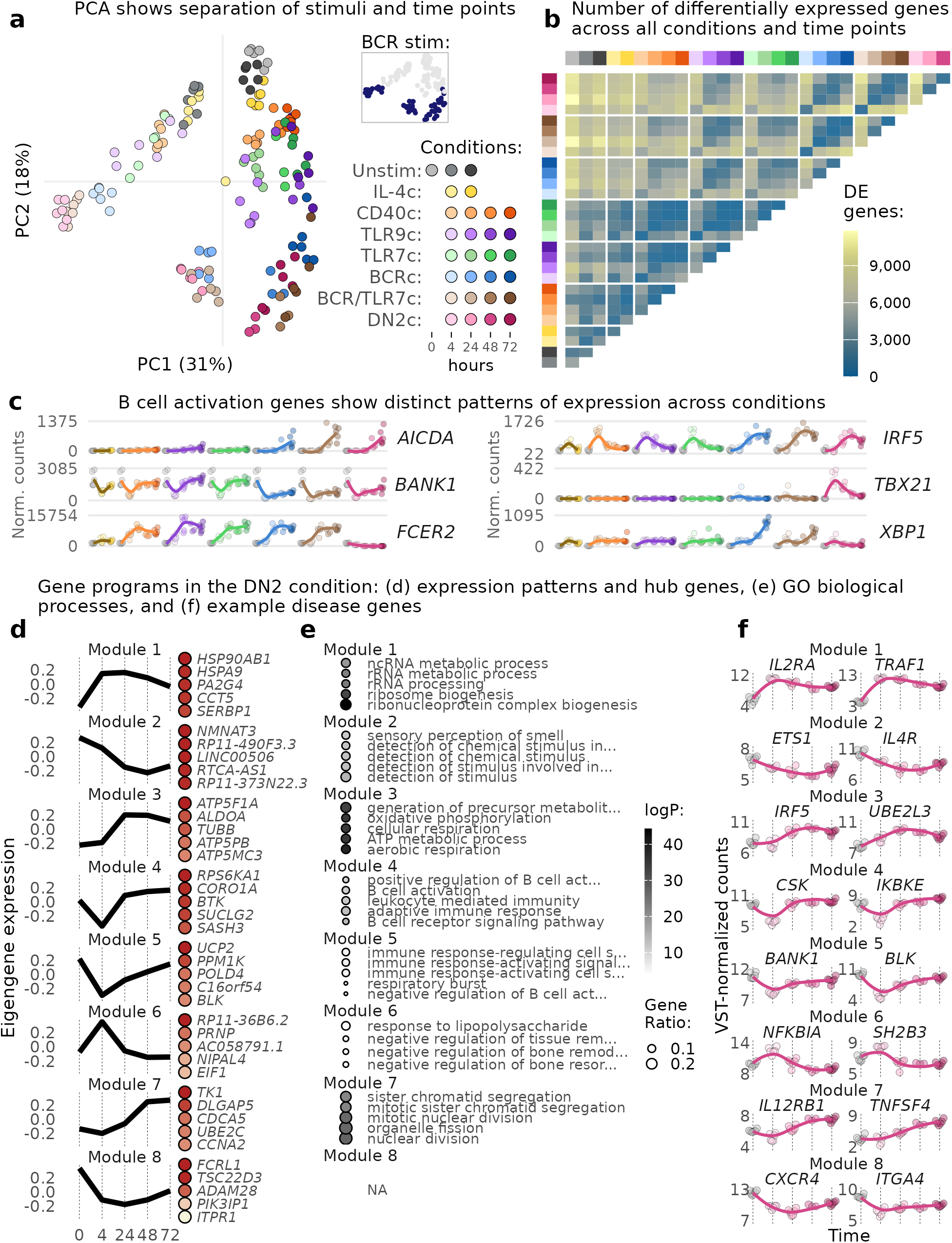
Low-input RNA-seq of 24 activation and 5 control conditions shows pathway-dependent gene expression and transcriptional dynamics in activated B cells over time. **a**, Principal component analysis (PCA) with the top 2,000 most variable genes shows separation of conditions and time points. Inset: PCA plot highlighting conditions that include BCR stimulation. **b**, Differential gene expression across all pairs of conditions and time points. **c**, Expression levels of selected B cell activation genes. X-axis represents time and colors represent conditions as in (a). **d**, Modules of gene expression identified by Weighted Gene Correlation Network Analysis (WGCNA). X-axis: time in hours. Y-axis: module eigengene expression values from WGCNA (eigengene is considered a representative of the gene expression profiles in a module). Main hub genes for each module are shown on the right-hand side of each panel, colored by kIM (WGCNA’s intra-module connectivity). **e**, Enrichment of gene ontology (GO) biological processes for modules in (d). **f**, Expression levels of selected IMD-associated genes belonging to modules in (d). Transcriptome-based clustering reveals 13 clusters (Figure 3b,d). Naive-like B cells expressing *TCL1A*, *FOS*, and *DUSP1* were mostly found among unstimulated cells (cluster 12). After stimulation, B cells committed to different fates. For example, cells from the BCRc at 72 hours are almost completely distributed into memory-like cells (cluster 4), early plasmablasts (cluster 0), and proliferating cells (clusters 9 and 10).

The RNA-seq profiling reveals stimulus-specific dynamic responses for genes following B cell stimulation (Figure 2c). For example, *AICDA*, which encodes the enzyme activation-induced cytidine deaminase involved in class switch recombination and somatic hypermutation, was strongly up-regulated in late time points of conditions that include BCR stimulation. *BANK1*, which encodes the B cell scaffold protein with ankyrin repeats 1 that transduces BCR signals, showed sustained down-regulation in conditions that include BCR stimulation, whereas expression quickly recovered after 4 hours in the other conditions. *FCER2* (which encodes CD23, an IgE receptor) was repressed only in the DN2-inducing condition (DN2c). *IRF5*, which codes for an interferon regulatory transcription factor with multiple functions, showed anti-correlated expression dynamics between conditions that include BCR stimulation and those that do not. *TBX21* (which encodes the transcription factor T-bet) was specifically activated in the DN2-inducing condition and not in any other condition. Finally, *XBP1*, which encodes X-box-binding Protein 1, a marker of plasma cell differentiation, showed early up-regulation at 4 hours in all conditions but has further strong up-regulation at 72 hours in BCRc, whereas in DN2c it was down-regulated.

Next, to understand the broad patterns of gene dynamics, we used weighted gene co-expression network analysis (WGCNA) to leverage the correlation of gene expression to group genes into transcriptional programs (“modules”).^24^ Here, we employed gene expression data from unstimulated B cells as the 0-hour baseline and transformed gene expression values using variance-stabilizing transformation (VST) to understand expression dynamics relative to time independently of total gene expression levels. When applied to the DN2c time course, we identified eight gene modules (Figure 2d). To understand the biological processes enriched in each module, we used Gene Ontology (GO) enrichment analysis (Figure 2e). This analysis unraveled the functional dynamics of B cell activation. For example, genes involved in B cell activation (modules 4, 5, and 8) are initially down-regulated with a rise in gene expression in late time points. Genes in module 4, which has *BTK* (necessary in BCR signaling) as one of its main hub genes, recover higher expression levels already at 24 hours, whereas genes in module 5, with *BLK* (also involved in BCR signaling) among the main hub genes, display a slower rise in expression levels. Module 8, with an even slower rise in expression levels, had no significant pathway enrichment, but its main hub gene is *FCRL1*, a promoter of B cell activation.²⁵ At the same time as these B cell activation genes are down-regulated, genes involved in RNA processing and ribosome biogenesis (module 1) go up and peak at 4 hours. At 24 hours, cellular respiration genes (module 3) are activated. We also see enrichment in module 3 for nucleoside synthesis and DNA unwinding related to duplication, suggesting that cells are preparing for division. Indeed, at 48 and 72 hours, genes involved in mitosis (module 7) are up-regulated. This “proliferation program” is observed only in the three conditions that include BCR stimulation. Overall, these enrichment patterns suggest that once B cells are activated, they de-prioritize transcription of B cell activation genes and prioritize transcription of translational machinery, which probably aids in subsequent production of energy which is then used for B cell proliferation —with the latter only if the BCR was stimulated.

This analysis enables us to assess the dynamic transcriptional programs to which IMD-associated genes belong (Figure 2f). As members of module 1 we have, for example, *IL2RA*, which encodes the alpha chain of the IL-2 receptor complex (also known as CD25) and is a central immune modulator associated with several IMDs,^26^–^36^ and *TRAF1*, whose expression drives risk of inflammatory arthritis in children and adults.^34^,^37^ *ETS1* and *IL4R* are members of module 2. *ETS1*, an important regulator of lymphocyte differentiation, is associated with multiple IMDs.^26,29^,^31^,^33^,^34^,^38^–^41^ *IL4R*, involved in IgE class switching and secretion from B cells among other functions,^4^² is associated with severe asthma and is a therapeutic target in asthma.^27^,^43^ As example genes in module 3, *IRF5* is an interferon response gene implicated in autoimmune diseases such as RA, SLE, Sjögren’s, and SSc,^34^,^40^,^44^–^46^ while *UBE2L3* is involved in ubiquitination to target proteins for degradation and is also associated with several IMDs.^31^,^32^,^34^,^40^,^46^,^47^ In module 4, *CSK* encodes a Src kinase that regulates B cell activation and is associated with SLE and SSc,^46^,^47^ and *IKBKE*, which is associated with SLE and encodes the kinase IKKε.^49^ In module 5, we have *BANK1* and *BLK*. *BANK1* is associated with SLE;^40^ while *BLK* is associated with RA, SLE, and SSc.^34^,^40^,^45^ In module 6, *SH2B3* encodes an adaptor protein involved in various signaling activities by growth factors and cytokine receptors, and is implicated in dozens of human traits including autoimmune diseases,^30^,^31^,^33^–^36^,^46^,^47^ and *NFKBIA*, a negative regulator of NFκB that is associated with asthma and psoriasis.^27^,^50^ In module 7, *IL12RB1* encodes a subunit of the IL-12 receptor that is associated with SSc,^4^⁸ whereas *TNFSF4* encodes OX40L on the surface of B cells and is associated with asthma, eczema, RA, and SLE.^27^,^46^,^51^,^52^ In module 8, *CXCR4* is associated with MS,^33^ and *ITGA4*, which encodes the integrin subunit alpha four involved in cell motility and signaling, and is associated with inflammatory bowel disease (IBD).^31^ Notably, within a disease, candidate causal genes may belong to different gene modules. For example, among B cells in the DN2c condition, four candidate SLE risk genes are tightly associated ( kME >= 0.9) with module 4, four genes with module 5, two genes with module 1, two genes with module 3, and one gene with module 7 (Figure S2). Overall, this wide dispersion of IMD–associated genes across gene modules underscores the complexity of IMDs, showing that candidate causal genes may play key roles at different time points of B cell response to activation.

### Single-cell profiling shows pathway-dependent B cell fates

We performed single-cell RNA-seq coupled with surface protein expression profiling (CITE-seq) on total B cells extracted from five genotyped healthy participants. We assayed the following conditions: unstimulated cells at day 0, and IL-4c, TLR7c, BCRc, and DN2c at 24 and 72 hours (Figure 1). We performed this experiment in three batches —one batch including two donors, and two batches on the same three donors— pooling cells from multiple conditions and donors into one run at each batch. Before pooling, cells from different conditions were tagged with specific hashtag oligonucleotides. After sequencing, demultiplexing, and quality control to remove empty droplets, doublets, and dying cells, we obtained 37,700 cells.

Single-cell RNA-seq revealed stimulus-dependent B cell states (Figure 3). By looking at both RNA expression and surface protein expression, we observed that BCR engagement in BCRc and DN2c leads to decreased expression of IgD/IgM on the surface of cells in comparison with the Day 0, IL-4c, and TLR7c conditions. At 24 hours, we also observed higher levels of CD69 protein expression in BCRc and DN2c in comparison to IL-4c and TLR7c, indicating high B cell activation after BCR stimulation (Figure 3c). The BCRc and DN2c conditions also showed the highest amounts of cells expressing *MKI67*, a marker of proliferation, and CD11c^+^ B cells. The BCRc condition led to the highest levels of IgD^-^ CD27^+^ B cells, a memory-like phenotype, and also cells expressing *XBP1*, a marker of plasma cell differentiation (Figure 3e).

**Figure 3:**
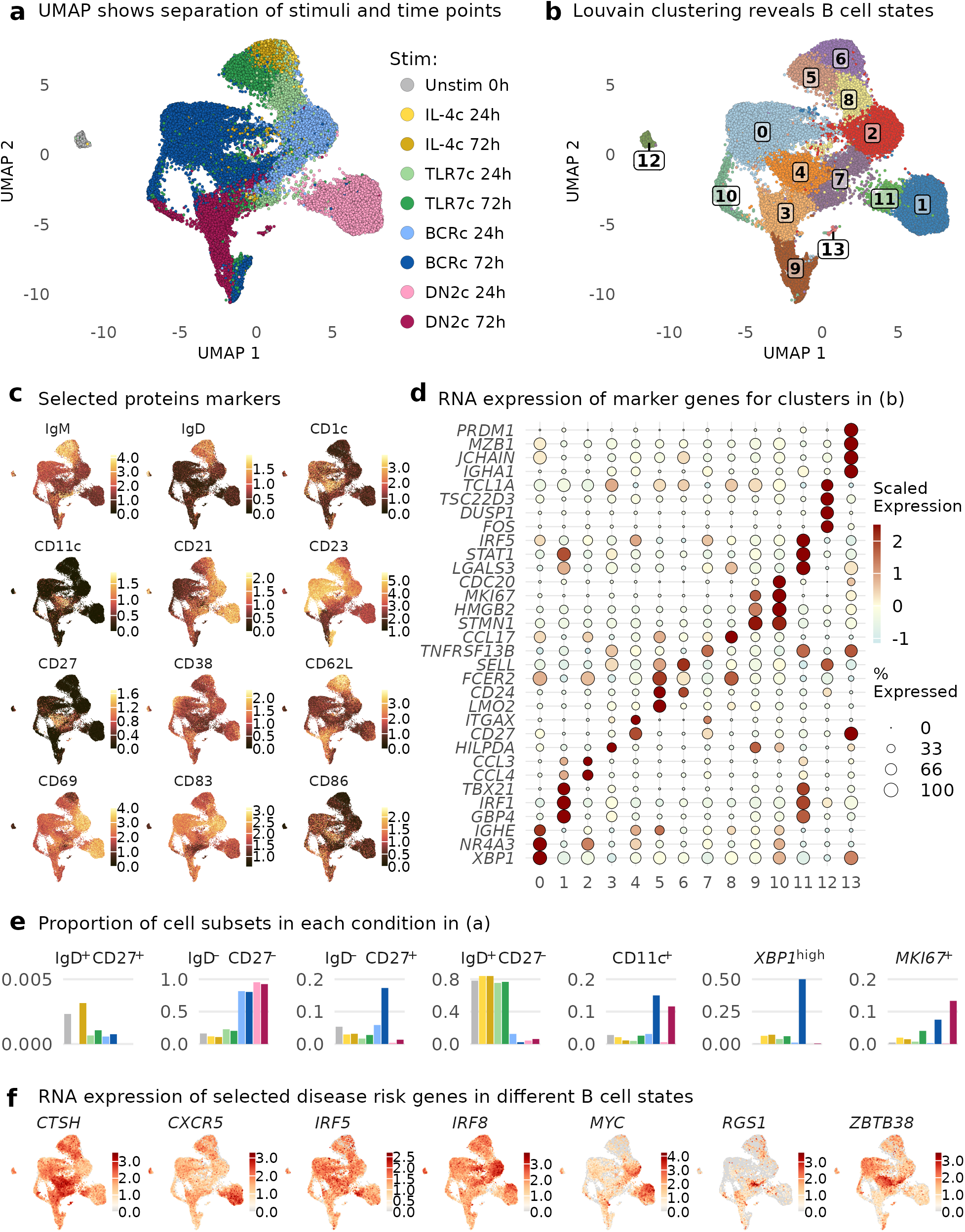
Single-cell RNA expression coupled with quantification of 137 surface protein markers reveals pathway-dependent B cell fates. **a**, Uniform Manifold Approximation and Projection (UMAP) based on gene expression after Harmony correction. **b**, UMAP as in (a) colored by transcriptome-based clusters. **c**, UMAP as in (a) colored by protein expression levels of selected surface markers. **d**, RNA expression levels of selected cluster marker genes. **e**, Proportion of B cell subsets defined by expression of marker protein/genes (genes in italic) within each condition and time point (colors as in (a)). **f**, UMAP as in (a) colored by RNA expression of selected IMD-associated genes.

Finally, we looked at the expression patterns of IMD-associated genes across different B cell states (Figure 3f). For example, *CTSH* —which encodes for Cathepsin H, is associated with MS and type 1 diabetes (T1D),^33^,^45^ increases at 72 hours after BCR stimulation— is mainly expressed in *CD27*-expressing cells. *CXCR5* encodes a B cell homing factor, it is in a risk locus for multiple IMDs,^27^,^33^,^53^ and in our data it is expressed within a subset of cells in the DN2c at 24 hours. *IRF5*, associated with many autoimmune diseases and one of the strongest non-MHC associations with SLE,^34^,^40^,^45^ is expressed mainly in cluster 11, a state of DN2c with high expression of *STAT1* and *LGALS3*. *IRF8*, another interferon response gene that is associated with IMDs such as IBD, PBC, SLE, and SSc,^31^,^46^,^54^–^56^ is more highly expressed in cells that co-express *MYC*, *CCL3*, and *CCL4*, in BCRc and DN2c at 24 hours. *RGS1*, which is associated with multiple autoimmune diseases,^33^,^47^,^57^,^58^ is expressed in BCRc at 72 hours, restricted to a small population expressing *ITGAX* and *FCRL5*. Finally, *ZBTB38*, associated with asthma,^27^ increases expression in the BCRc condition with time, consistent with the bulk RNA-seq data. The data at single-cell resolution reveals that such an increase is driven by cells co-expressing *ITGAX* and *FCRL5*.

### B cell activation induces epigenomic changes relevant for IMDs

In addition to gene expression, we used ATAC-seq on stimulated B cells to gain insight into how chromatin is remodeled following stimulation (Figure 4). We performed ATAC-seq on three healthy donors for conditions IL-4c, TLR7c, BCRc, and DN2c, at 24 hours, and used unstimulated cells at 0 hours and in media only for 24 hours as controls.

**Figure 4:**
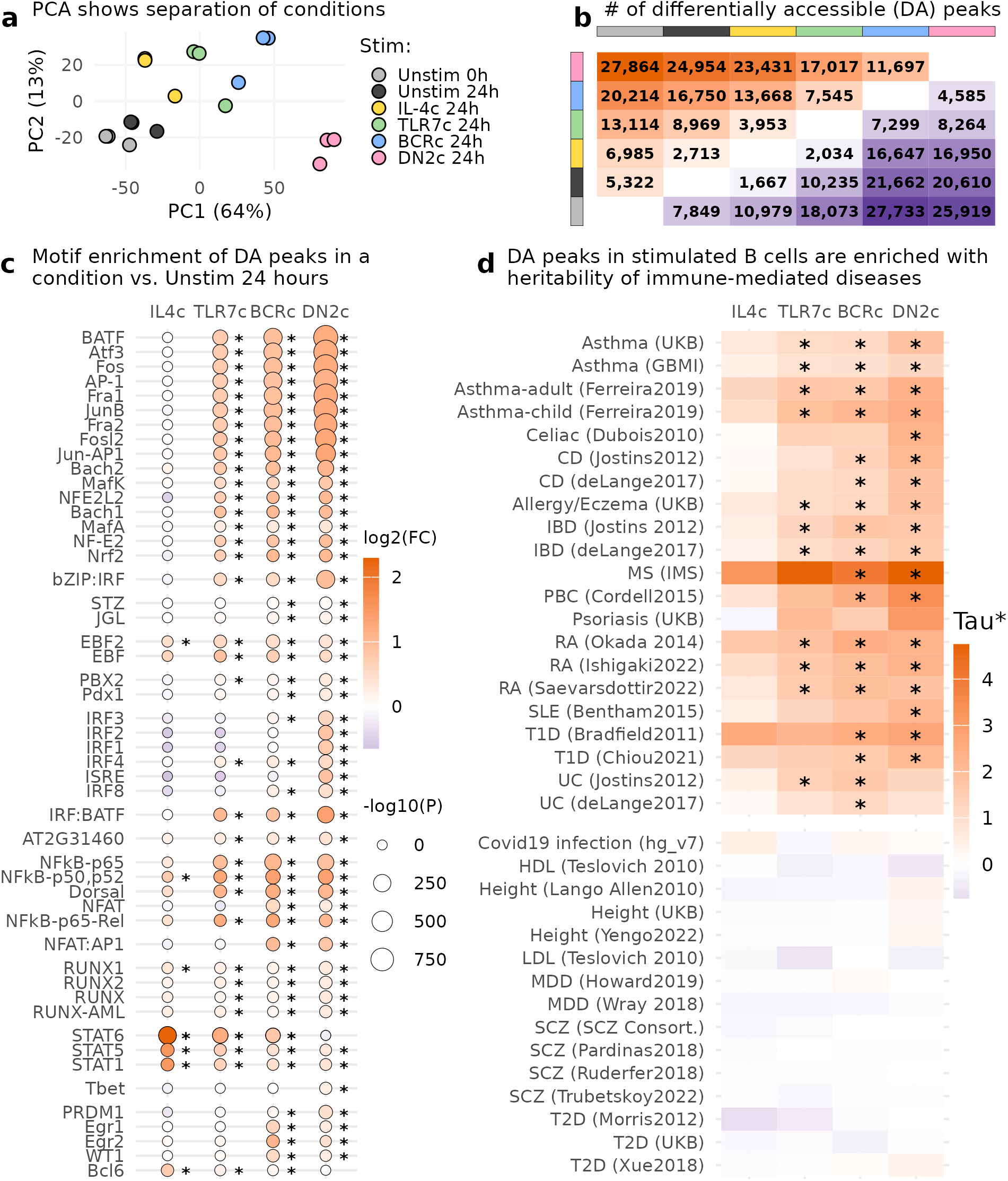
Open chromatin profiling in activated B cells using bulk ATAC-seq uncovers disease-relevant cell states. **a**, Principal component analysis on the 5,000 most variable peaks. **b**, Pairwise number of differentially accessible peaks across all conditions (1% FDR). In orange, peaks that are more accessible in condition in the rows in comparison to condition in the columns. In purple, peaks that are less accessible in condition in the columns in comparison to conditions in the rows. **c**, Transcription factor motif enrichment in differentially accessible chromatin regions in stimulated conditions with respect to unstimulated cells at 24 hours. Colors indicate log₂ fold change, and size indicates -log₁₀ p-values from HOMER. **d**, Enrichment of heritability for IMDs (top panel) and control traits (bottom panel) in differentially accessible chromatin regions in stimulated conditions with respect to unstimulated cells at 24 hours, estimated with LDSC-SEG. Tau*: normalized effect size. Asterisks indicate significance at 1% FDR.

We observed significant differences in chromatin accessibility across conditions. We performed PCA on the top 5,000 most variable peaks. PC1 captures a gradient of activation strength, with control conditions on the left, followed by TLR7c, then BCRc, and finally DN2c at the other extreme (Figure 4a). We performed differential accessibility analysis across all pairwise combinations of conditions (Figure 4b). The number of differentially accessible peaks between activation conditions and unstimulated cells goes in line with the observed PC1 gradient, with DN2c presenting the highest number of differentially accessible regions compared to unstimulated cells at 24 hours, followed by BCRc and then TLR7c. These results are concordant with what we observed in the RNA-seq data, with DN2c being the strongest stimulation, inducing the largest number of changes.

To identify which transcription factors might be involved in the chromatin remodeling of B cells after activation, we selected peaks that are differentially more accessible at 1% FDR in an activation condition compared to the unstimulated condition at 24 hours (second column in Figure 4b). We further filtered for peaks with a large effect size (log₂ fold change > 1) and employed HOMER to identify transcription factor (TF) motifs enriched in open chromatin regions at each activation condition (Figure 4c).⁵^9^ Overall, activated B cells demonstrated enrichment of multiple motifs, with the DN2c condition exhibiting the highest number of enriched motifs and the greatest fold enrichment. Motifs of TFs that harbor basic leucine zipper (bZIP) domains showed the strongest enrichments, with the most significant TF motif being the one of BATF. BATF has previously been described as a pioneer transcription factor involved in chromatin remodeling and class switch recombination in lymphocytes.^60^,^61^ The DN2c condition also exhibited significant enrichment of the IRF family of TFs, as well as *PRDM1* (BLIMP-1), and T-bet.

Finally, we used linkage disequilibrium score regression for specifically expressed genes (LDSC-SEG) to integrate our chromatin data with GWAS data to test whether chromatin regions that become accessible following stimulation are enriched for genetic risk of IMD. This is a method that has been widely used to find the specific cell types or cell states at which disease risk variants likely affect gene regulation, hence pointing to key cell states that mediate genetic susceptibility (e.g., activated T cells for RA, rhinovirus-infected airway epithelial cells for asthma).^19^,^62^,^63^ We utilized this method by providing annotations composed of the open chromatin regions that are significantly more accessible in each of the activation conditions at 24 hours versus the 24 hour non-stimulated control (1% FDR), i.e. IL-4c vs. non-stim, TLR7c vs. non-stim, BCRc vs. non-stim, and DN2c vs. non-stim. We then provided summary statistics of 15 immune-mediated diseases (some of which had multiple independent GWAS performed), and 6 “negative control” diseases or traits for which we do not expect an immune-mediated mechanism, hence we do not expect an enrichment of genetic risk in open chromatin regions specific for activated B cells. Using this method, we observed significant heritability enrichment (1% FDR) in activation-dependent peaks for multiple IMD traits, including RA, SLE, T1D, MS, PBC, IBD and its subtype CD, and celiac disease (Figure 4d). For most of these diseases, the heritability enrichment was higher for the open chromatin regions that become more accessible with the DN2c. To note, we did not find significant enrichment in “negative control” traits for which we would not expect activated B cells to be relevant, such as type 2 diabetes (T2D), schizophrenia (SCZ), major depressive disorder (MDD), LDL and HDL cholesterol, and height (Figure 4d). We included a COVID-19 infection GWAS given that it has been reported that patients with severe COVID-19 present expansion of DN2 B cells,^64^ but we did not find a significant heritability enrichment in open chromatin regions specific for activated B cells for this trait. Overall, these findings underscore the relevance of studying activated B cells to understand genetic associations with IMDs and suggest that some GWAS associations with yet unexplained mechanisms can be uncovered by studying their regulatory effects in activated B cells.

### B-cell activation-specific chromatin region reveals function of OX40L variant in immune diseases

Motivated by the IMD heritability enrichment in stimulated B cells, we sought to investigate whether open chromatin regions of activated B cells could provide insights into the function of specific risk variants. To do this, we fine-mapped SLE GWAS data and intersected it with our ATAC-seq data to find putative causal variants falling in open chromatin regions that are specific to activated B cells. As proof of concept for this approach, we decided to focus on the 1q25.1 risk locus, where a single candidate causal variant is found after fine-mapping. Manku et al. performed a trans-ancestral association study at this locus and found that rs2205960 consistently explains SLE risk in different ancestries.^65^ Additionally, we performed fine-mapping with in-sample linkage disequilibrium estimation in the Langefeld et al. GWAS data.^40^ Rs2205960 is the top variant at the 1q25.1 locus, and was the single variant included in the main credible set with a posterior probability greater than 0.99 (Figure 5a).

**Figure 5:**
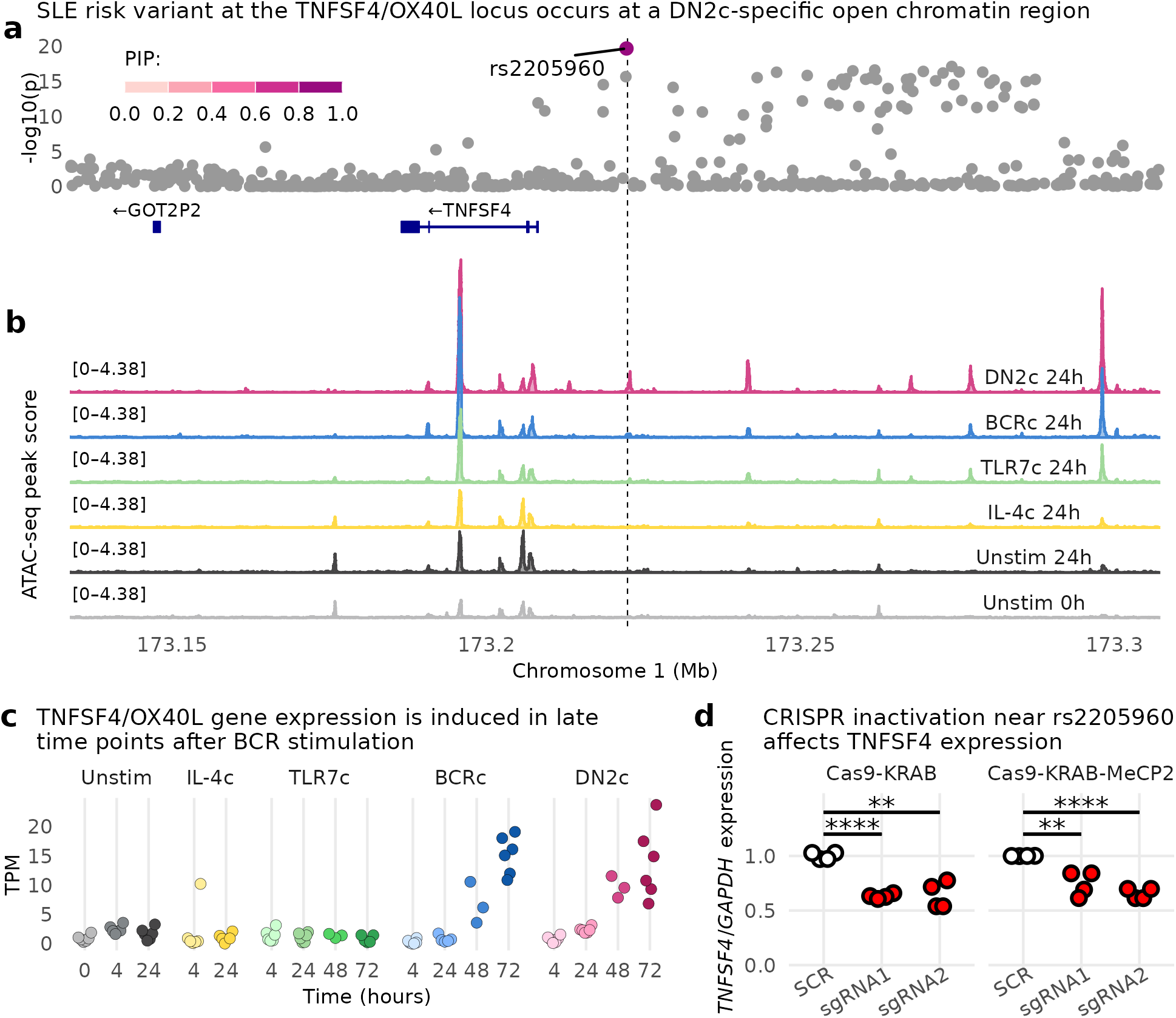
SLE risk variant overlapping DN2c-specific open chromatin region is in an enhancer that regulates *TNFSF4*. **a**, GWAS data from Langefeld et al.^40^ rs2205960 is the most likely causal variant. PIP: posterior inclusion probability from SuSiE.^66^ **b**, Chromatin peaks at the genomic region in (a). **c**, *TNFSF4* gene expression in the same conditions shown in (b). TPM: transcripts per million. **d**, CRISPRi shows that inhibiting the region containing the putative causal variant leads to down-regulation of *TNFSF4* RNA expression.

This intergenic variant is located 15kb 5′ of *TNFSF4*, within a region where, in our data, chromatin is open exclusively in the DN2c condition (Figure 5b). *TNFSF4* is expressed at late time points in conditions that include BCR engagement (BCRc and DN2c) and is more expressed in the DN2c condition than in BCRc (Figure 5c). To assess whether the open chromatin region overlapping rs2205960 functions as a regulatory element for *TNFSF4* expression, we used CRISPR inactivation (CRISPRi) in the lymphoblastoid cell line (LCL) GM12878. This Epstein-Barr Virus (EBV)-transformed B cell line model constitutively activated B cells. First, we generated stable LCLs dCas9-KRAB and dCas9-KRAB-MECP2 systems, which have repressor domains that can epigenetically suppress the target region of interest. We designed two single guide RNAs to bind near rs2205960 and we nucleofected them separately in each of the CRISPRi system cell lines. We observed that inhibiting the region that contains the variant leads to reduced *TNFSF4* expression regardless of single guide RNA or cell line used (Figure 5d), demonstrating this region is an enhancer that regulates *TNFSF4*.

### Pervasive stimulus-dependent splicing changes during B cell activation

Cell states are shaped not only by the epigenome and the level of expression of genes, but also by what particular isoforms are expressed by each gene. To quantify alternative splicing in a robust way, high-depth RNA-seq libraries that cover the full transcript are needed. Low input RNA-seq yields full transcript information but, starting from 1000 cells per sample, it yields low complexity libraries that present shortcomings for isoform quantification. Droplet-based single-cell RNA-seq only quantifies one end (5′ or 3′) of each gene, so which middle exons get included or excluded is not possible to ascertain. Hence, we performed standard RNA-seq with deep sequencing (at least 40M pairs of 150bp reads per sample) in 16 healthy donors. We utilized 500,000 cells per condition, to capture a wide dynamic range of expressed isoforms. Given the larger input material, we performed full transcriptome profiling on four conditions only per donor: untouched B cells (considered the 0 hour time point), as well as TLR7c, BCRc, and DN2c for 24 hours (Figure 6a).

**Figure 6:**
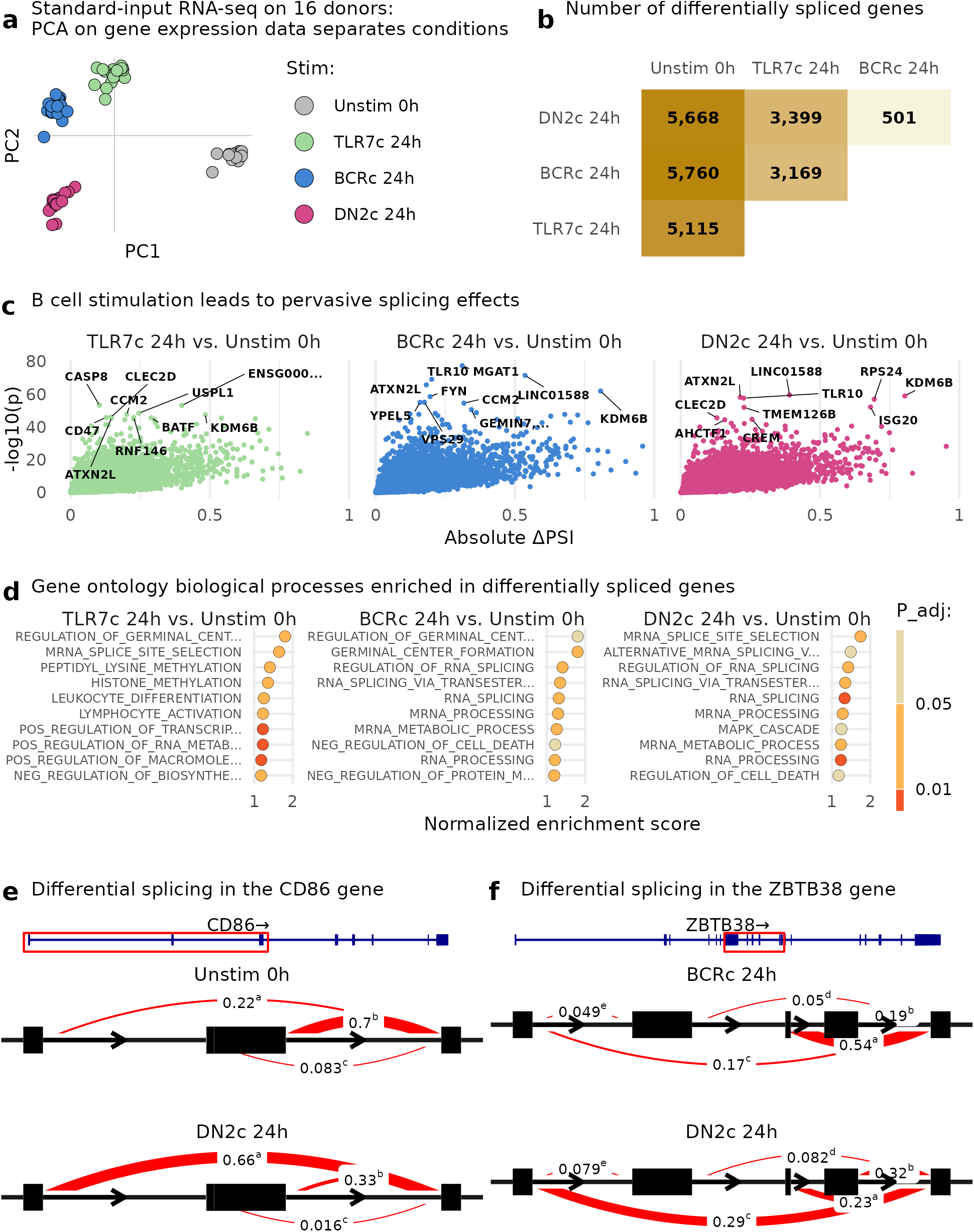
Pervasive alternative splicing changes during B cell activation. **a**, Principal component analysis on gene expression data from 16 donors. **b**, Number of differentially spliced genes at 5% FDR between all pairs of conditions. **c**, Splicing effect size in ΔPSI (percent spliced in) between each activation condition and un-stim by -log₁₀ of the p-value. **d**, Gene ontology biological processes enriched in differentially spliced genes in (c). P-value and enrichment scores from the fgsea R package. **e**, Differential splicing event in the *CD86* gene between DN2c and un-stim. **f**, Differential splicing event in the *ZBTB38* gene between BCRc and DN2c. Numbers indicate average proportion of junction usage, and linewidths are scaled to this usage.

Next, we quantified RNA splicing with the LeafCutter pipeline,^67^ which uses an annotation-free algorithm to measure intron excision rates and performs differential splicing analysis with a Dirichlet-multinomial model. At 5% FDR, we identified 5,115, 5,760 and 5,668 genes with differential splicing between unstimulated B cells and 24 hour-activated B cells with TLR7c, BCRc, and DN2c, respectively (Figure 6b,c). Between activation conditions, we found 501–3,399 genes with significant differential splicing at 5% FDR, with the largest differences found between TLR7c and the two conditions that simulate the BCR (BCRc and DN2c, Figure 6b). We performed pathway enrichment analysis on the differentially spliced genes between each of the activation conditions and non-stimulated cells (Figure 6d). For all three activation conditions “mRNA splice site selection” was within the top 2 pathways enriched, with other pathways related to RNA splicing also significantly enriched. This has also been observed in activated T cells and PBMCs, where genes that code for proteins involved in alternative splicing and/or RNA processing, such as *CELF2*, *HNRNPD*, and *SFRS2*, present differential splicing upon immune activation.^68^ To note, differentially spliced genes for TLR7c and BCRc are enriched in germinal center formation pathways, but this pathway is not significant in DN2c. In Figure 6e,f we show examples of genes that present stimulus-dependent differential splicing. We observe that in *CD86*, an activation marker, splicing excludes exon 2 more often in DN2c than in non-stimulated cells. In *ZBTB38*, an asthma risk gene,^27^ splicing leads to a different exon composition between BCRc and DN2c. Overall, our analysis of alternative splicing shows that isoform usage drastically changes in B cell activation, and which specific activation pathways are induced can also influence alternative splicing of genes.

## Discussion

B cells play crucial roles in our immune system, such as producing antibodies and modulating other immune cell types. However, their dysfunction is implicated in many IMDs. Despite their importance in health and disease, the precise mechanisms by which disease-associated genes and genetic variants influence B cell activation and disease remain poorly understood. In this study, we generated multiomics data to comprehensively profile gene expression and chromatin remodeling during B cell activation through multiple receptor pathways.

Using bulk RNA-seq, we profiled how B cells transcriptionally respond to different activation conditions over time. We identified gene programs (or modules) that illuminate how B cells respond to stimulatory signals, including transcriptional changes associated with B cell activation, energy generation, ribosome biogenesis, protein translation, and cell division. Each of these modules exhibits distinct temporal dynamics. While these modules are conserved across activation conditions, BCR stimulation is crucial for the activation of a “proliferation program” at 48 and 72 hours. This program is observed in all three conditions that include BCR stimulation, and it is most prominent in DN2c. For example, the gene module itself is larger in the DN2c condition. Further, gene ontology analysis of genes overexpressed in DN2c compared to BCRc at 72 hours reveals significant enrichment for biological processes related to DNA replication, chromosome segregation, and nuclear division (Table S1). In conditions lacking BCR stimulation (CD40c, TLR7c, TLR9c), the proliferation program is not detected. Additionally, in non-BCR stimulated conditions, a module of genes coding for ribosomal proteins, strongly enriched for “cytoplasmic translation”, decreases in expression over time. Notably, this down-regulation does not occur upon BCR stimulation, indicating that BCR engagement might sustain protein synthesis for longer time periods (Figure S3, Figure S4, Figure S5, Figure S6, Figure S7).

To investigate B cell activation at the single-cell level, we performed single-cell RNA-seq. We found that B cells follow different cell fates depending on the activation condition. By applying clustering and differential gene expression analysis, we identified naive B cells, memory-like B cells, *XBP1*-expressing B cells, and proliferating B cells, and how the frequency of these subsets vary depending on the experimental condition. After BCR stimulation in BCRc and DN2c, IgM/IgD protein expression is markedly reduced by 24 hours, and by 72 hours, a proliferating *MKI67*^+^ subset emerges, with this subset being more abundant in DN2c. These results concord with the observations of the proliferation gene module identified in the bulk RNA-seq analysis. Furthermore, the majority of CD27^hi^ cells at 72 hours originate in BCRc, whereas DN2c appears to suppress this cell state.

Next, we performed ATAC-seq to study the chromatin remodeling at 24 hours post-stimulation. Our data suggest a gradient of activation strength across conditions: IL-4c, TLR7c, BCRc, and DN2c, with DN2c exhibiting the most robust activation. Transcription factor motif analysis of activation-dependent open chromatin regions revealed strong enrichment for bZIP-containing TFs, such as BATF and ATF3, in all conditions except IL-4c, with enrichment increasing toward DN2c. On the other hand, binding sites of the transcriptional repressor BCL6 show the opposite trend, with no significant enrichment in DN2c. Notably, DN2c exhibits specific or enhanced motif enrichment for IRF family TFs (IRF1, IRF2, IRF3, IRF4, IRF8), BLIMP-1 (*PRDM1*), and T-bet (*TBX21*). Most of these TFs are in risk loci for IMDs. For example, *IRF8* is a likely target gene for risk variants for SLE, UC, PBC, MS, SSc, and RA.^31^,^33^,^34^,^46^,^48^,^53^,^54^,^56^ *PRDM1* is a likely target gene for variants associated with IBD, UC, CD, and psoriasis.^31^,^39^,^55^,^69^ *TBX21* is in risk loci for autoimmune thyroid disease, asthma, childhood-onset asthma, and eczema.^27^,^30^,^70^ These findings suggest that studying human B cells activated with the DN2c can be of high relevance to understand the role of IMD-associated TFs in B cell function and differentiation.

By integrating our chromatin data with publicly available GWAS data, we found that activation-dependent open chromatin regions are enriched for immune-mediated disease heritability. Although this enrichment does not necessarily imply causality of risk variants in B cells, it suggests that studying risk variants in activated rather than resting B cells could provide valuable insights into IMD pathogenesis. As a proof of concept, we examined the rs2205960 SLE risk variant at the *TNFSF4* locus.^40^ *TNFSF4* encodes OX40L, a tumor necrosis factor (TNF) superfamily member expressed on the surface of antigen-presenting cells that binds OX40 (*TNFRSF4*) on the surface of T cells to activate NFκB. The *TNFSF4* locus is associated with other immune-mediated diseases apart from SLE, such as RA, asthma, and eczema.^27^,^46^,^51^,^52^ OX40L–CAR-Tregs have been developed to deplete OX40L-expressing cells and control alloreactivity and autoimmunity.^71^ Our data shows that *TNFSF4* is lowly expressed in resting B cells, but it is strongly up-regulated upon BCR stimulation (in BCRc and DN2c), thus suggesting that activation is required for exhibiting its functions. Further, we show that the risk variant is located in a genomic region with accessible chromatin specifically in the DN2c condition, highlighting the importance of selecting appropriate activation states when studying the functional impact of risk variants.

Lastly, we explored splicing changes during B cell activation. Splicing has gained attention in cancer research, leading to the development of therapeutic splicing modulators.^72^ Moreover, recent studies have shown that a proportion of the IMD risk variants (17%–25%) might exert their effects by altering RNA splicing.^73^,^74^ For example, Mu et al. reported that among sixteen SLE GWAS loci that co-localized with different types of QTLs, ten co-localized exclusively with splicing QTLs.^73^ We propose that investigating splicing across diverse B activation states may help elucidate uncharacterized IMD associations. Here we provide a comprehensive catalog of splicing changes occurring during B cell activation. This resource will aid in the identification of splicing-mediated regulatory mechanisms and support the functional interpretation of IMD-associated variants.

Despite the broad range of stimuli and high depth of molecular profiling in our study, a number of limitations should be considered. First, the study was performed using *in vitro* stimulation of B cells from healthy adult female donors. While this design facilitates controlled perturbation and avoids disease-related confounders, it may not fully capture the *in vivo* complexity of B cell activation in disease contexts or across diverse populations of different age groups, sex, and genetic ancestries. Second, although we incorporated a wide spectrum of stimuli relevant to key immunological pathways, additional microenvironmental cues that exist in tissue niches (such as particular cytokine combinations and interactions with other cell types) are not captured in this system. Furthermore, it is hard to know whether the concentration of our stimulants is lower or higher than that encountered by B cells in the human body. Third, while we linked open chromatin regions to disease-associated variants, further experimental validation will be needed to confirm the causal variants and their precise gene targets.

Our work is complementary to previous studies of B cell multi-omics. For example, Guo et al. conducted bulk RNA-sequencing, proteomics and metabolomics analyses on B cells cultured for 24 hours in control and activation conditions (10 in total), many of which similar to those used in our study.^75^ This overlap allows for direct comparisons while also enabling us to expand upon their findings using our additional time points, ATAC-seq and single-cell RNA-seq data. For instance, the authors found that *BCAT1* is highly up-regulated following combined simulation of BCR and TLR9, leading to mTOR activation and promoting B cell growth. In our dataset, *BCAT1* exhibits stronger activation in the DN2c condition. Our multiple time points reveal that *BCAT1* is associated with module 1 (Fig.2), a gene module characterized by early activation at 4 hours.

In summary, our multi-omics approach provides a comprehensive profile of B cell activation in response to different stimuli. This publicly available and easy-to-access resource will facilitate future studies investigating B cell function and its implications in immune-mediated diseases.

## Methods

### Samples

We recruited 26 adult female donors aged 20–60 years from the Mass General Brigham Biobank (MGBB) through the Joint Biology Consortium Recruitment Core, and the Boston Children’s Hospital Biorepository of Adult Healthy Controls. All donors were recruited as part of lupus research studies.

### B cell processing

Blood samples were collected from healthy donors using EDTA-coated tubes. Peripheral blood mononuclear cells (PBMCs) were subsequently isolated by Ficoll-Paque (GE Healthcare) density gradient centrifugation. B cells were further isolated from PBMCs using a Total B Cell Isolation Kit (Stemcell Technologies), and their purity was confirmed through flow cytometry (FACS) analysis. Cells have been stimulated with the following conditions for the different experiments: **1) IL-4c:** 20ng/ml IL-4; **2) CD40c:** 20ng/ml IL-4 + 50ng/ml sCD40L; **3) TLR9c:** 20ng/ml IL-4 + 2.5μg/ml CpG; **4) TLR7c:** 20ng/ml IL-4 + 1μM R848, **5) BCRc:** 20ng/ml IL-4 + 10μg/ml anti-IgG/IgM, **6) BCR/TRL7c:** 20ng/ml IL-4 + 10μg/ml anti-IgG/ IgM + 1μM R848; **7) DN2c:** 10ng/ml BAFF + 10ng/ml IL-21 + 2.12ng/ml IL-2 + 20ng/ml IFN-y + 1μg/ml R848 + 5μg/ml anti-IgG/IgM.

### Low-input bulk RNA sequencing

Total B cells were activated as previously described. At the designated time points, live B cells were sorted directly into lysis buffer. RNA-seq for total B cells from six participants was performed at the Broad Institute using the SmartSeq-v2 library preparation and 75bp paired-end reads, including two control conditions and six activation conditions (Figure 1). Samples with less than 2 million sequenced reads were removed. We trimmed adapter sequences from the reads and performed QC with *trim_galore* and FastQC.^76^,^77^ We aligned reads to the reference transcriptome GRCh38 with Salmon (v1.5.1).^78^

We performed PCA in R (v4.1.2) with the top 2,000 most variable genes after VST normalization with DESeq2 (v1.34).^79^ In order to test for differential gene expression across conditions and time points we used edgeR v3.36,^80^ applying default filtering of minimum expression levels. We use a significance threshold of p < 0.05 on the Benjamini & Hochberg adjusted p-values. We used WGCNA v1.72 to leverage correlation of gene expression to identify gene modules.^24^ We used genes with at least 4 reads per million in at least 3 samples to build a signed network using Pearson correlation, a minimum module size of 60 genes, and merged closely related modules at a merge cut height of 0.2. To understand the biological processes enriched in each module, we performed gene ontology (GO) analysis with the clusterProfiler package (v4.2.2),^81^ after selecting genes with kME >= 0.9 in any module and the top 500 genes in each module by kME value.

### Standard-input bulk RNA sequencing

We performed RNA-seq on samples from 16 participants, at resting state and three stimulation conditions, targeting 40 million reads. Total B cells were activated under the specified conditions and time points. Following culture, cells were resuspended in lysis buffer, and RNA was extracted using the AllPrep DNA/ RNA Mini or Micro Kit (Qiagen), depending on the cell number. We performed QC with trim_galore and FastQC.^76^,^77^ We aligned reads to the reference genome GRCh38 with STAR (v2.7.9a) in two-pass mode, accepting up to 4% of mismatches, and adding the WASP tag.^82^ In order to identify possible sample swaps we used the MBV method to evaluate the concordance of the RNA-sequencing reads with the genotypes from MGB Biobank.^83^

### Single-cell RNA sequencing

B cells from five healthy donors were activated using a reverse time course to accommodate simultaneous processing and staining. Cells were cultured and activated for 72 hours, 24 hours, or left unstimulated (0h). At the 0h time point, all cells were collected and stained with Hash Tag Oligonucleotides (HTOs) and Antibody-Derived Tags (ADTs) using TotalSeq^TM^-C antibodies (BioLegend). The samples were then pooled, and live (7-AAD-) CD19^+^ cells were sorted for sequencing. Droplet encapsulation and sequencing were performed at the Center for Cellular Profiling, Brigham and Women’s Hospital, using the droplet-based 10X Genomics Chromium scRNA-seq platform. Sequencing was performed in two batches.

We used Seurat v4 for quality control and data analysis.^84^–^87^ We removed droplets with less than 500 genes and more than 10% of mitochondrial reads. We performed HTO-based demultiplexing with demuxmix v1.6.0,^88^ and genotype-based demultiplexing with demuxlet.⁸^9^ We integrated data from different libraries and donors with Harmony v1.2.0.^90^ We performed clustering and differential gene expression with standard Seurat functions.

### Bulk ATAC-seq

We isolated 100,000 nuclei from B cells and delivered them to the Center for Functional Cancer Epigenetics, Dana-Farber Cancer Institute, for ATAC-seq. Sequencing data was processed with the Nextflow ATAC-seq pipeline v2.0.^91^,^92^ Differential accessibility analysis was performed with DESeq2 v1.34.^79^ Differentially expressed peaks at 1% FDR in an activation condition at 24 hours with respect to the unstimulated condition at 24 hours were selected for downstream analyses. We used HOMER v4.11 for motif enrichment analysis.^59^ For heritability enrichment analysis, we used the linkage disequilibrium score regression (LDSC) method, with pre-processed GWAS summary statistics, linkage disequilibrium values, and SNP lists made available by the authors.^62^,^93^ The regression coefficients were normalized to Tau* as performed in (ref^94^) to facilitate comparison across traits.

### CRISPR validation experiments

GM12878 lymphoblastoid cell line (LCL), an ENCODE Tier 1 EBV-transformed human B cell line, was obtained from the Coriell Institute. Cells were cultured in RPMI 1640 medium supplemented with 10% fetal bovine serum, 1% penicillin-streptomycin, and 1% L-glutamine, at a density of 1 × 10⁵ cells/mL at 37°C in a humidified incubator with 5% CO₂.

To generate GM12878 cells stably expressing the CRISPR interference (CRISPRi) system, we produced lentiviral particles using either the lentiCRISPRi(v1)-Blast plasmid (Addgene #170067, expressing dCas9-KRAB) or lentiCRISPRi(v2)-Blast, according to previously published protocols.^9^⁵ Two single guide RNAs (sgRNAs) targeting the regulatory region containing the rs2205960 variant were designed using the CRISPRpick tool (Broad Institute), selecting guides with the highest predicted on-target efficacy and minimal off-target potential. The sgRNA sequences were as follows:

sgRNA1: AATAAAGCCTGACTAAGTAA

sgRNA2: ACCACACACTGGTGATCTAT

Non-targeting scrambled sgRNA (negative control): GCACTACCAGAGCTAACTCA

Nucleofection was performed using the SE Cell Line 4D-Nucleofector^®^ X Kit (Lonza) following the manufacturer’s instructions. Briefly, 300,000–500,000 GM12878 cells were resuspended in SE solution and nucleofected with 6µL of 30µM sgRNA using program DN-100. Immediately after nucleofection, cells were transferred into pre-warmed R10 medium and cultured for 72 hours to allow gene silencing.

Total RNA was extracted using a micro-RNA isolation kit (Qiagen), followed by cDNA synthesis using AffinityScript cDNA Synthesis Kit (Agilent). Quantitative PCR (qPCR) was performed using SYBR Green Master Mix on a QuantStudio 3 Real-Time PCR System (Applied Biosystems). *TNFSF4* transcript levels were measured using the following primers:

*TNFSF4* forward primer: CCTACATCTGCCTGCACTTCTC

*TNFSF4* reverse primer: TGATGACTGAGTTGTTCTGCACC

Housekeeping gene *GAPDH* forward primer: CCACATCGCTCAGACACCAT

*GAPDH* reverse primer: GGCAACAATATCCACTTTACCAGAGT

Data are presented as relative gene expression normalized to the housekeeping gene *GAPDH*, using the ΔΔCt method.

### GWAS fine-mapping

We performed statistical fine-mapping of the Langefeld et al. SLE GWAS data.^40^ We selected bi-allelic SNPs that were genotyped in the immunochip in a window of 500kb at each genome-wide significant locus, and applied SuSiE with in-sample LD to estimate variant probabilities and credible sets with at least 90% coverage.^66^

### Splicing

For splicing analyses, we have mapped the reads as in *Methods: Standard-input bulk RNA sequencing*, but adding the flag “--outSAMstrandField intronMotif” for STAR. We used regtools v0.5.1 to extract junctions.^96^ We performed intron clustering and differential splicing analysis between pairs of conditions with leafcutter v0.2.9.^67^ For intron clustering, we required that a junction needed to contribute at least 1% of the total reads within an intron cluster, and for differential splicing we used default parameters. We performed enrichment analysis on differentially spliced genes with the fgsea R package v1.20.0,^97^ ranking genes by their most extreme p-value across intron clusters, and testing enrichment in Gene Ontology biological processes. We used the leafviz R package for visualization of splicing events (https://github. com/jackhump/leafviz).

## Data Availability

All the sequencing data generated in this study will be made available in GEO and dbGAP. We made processed data publicly available via an interactive website that can be easily queried for visualization of gene expression, single cell profiles, chromatin accessibility, and splicing events: https://mgalab. shinyapps.io/bcellactivation.

## Code availability

Code is available at https://github.com/gutierrez-arcelus-lab/bcellactivation. Most software used in this project was installed and configured by Biogrids.^9^⁸

## Ethical approval

The Mass General Brigham and Boston Children’s Hospital Institutional Review Boards gave ethical approval for this study.

## Acknowledgments

We would like to thank Dr. Duane Wesemann and his laboratory (Brigham & Women’s Hospital) for helpful discussions on B cell activation, Stephen Gazal for guidance on application of LDSC-SEG, the Gutierrez-Arcelus and Nigrovic laboratories for helpful comments.

Vitor R. C. Aguiar was supported by a Joint Biology Consortium microgrant (Parent grant NIH/NIAMS 2P30 AR070253). Maria Gutierrez-Arcelus was supported by P30AR070253, the Lupus Research Alliance Lupus Innovation Award, the Lupus Research Alliance Empowering Research Career Development Award, the Vic Braden Family Arthritis National Research Foundation fellowship, the Gilead Sciences Rheumatology Research Scholars Award. This work was supported by two Boston Children’s Hospital Office of Faculty Development/Basic & Clinical Translational Research Executive Committees Faculty Career Development Fellowships (to Vitor R. C. Aguiar and Maria Gutierrez-Arcelus). Nicolaj Hackert was supported by an MD fellowship from the Boehringer Ingelheim Fonds. Peter A. Nigrovic was supported by the Lupus Research Alliance, NIH/NIAMS 2P30AR070253, 2R01AR065538, R01AR073201, and R01AR075906. Jeffrey A. Sparks is supported by the National Institute of Arthritis and Musculoskeletal and Skin Diseases (grant numbers R01 AR080659, R01 AR077607, P30 AR070253, and P30 AR072577), the National Heart, Lung, and Blood Institute (grant number R01 HL155522), the Rheumatology Research Foundation, the Arthritis Foundation, the R. Bruce and Joan M. Mickey Research Scholar Fund, and the Llura Gund Award funded by the Gordon and Llura Gund Foundation. The project described was supported by Clinical Translational Science Award 1UL1TR002541-01 to Harvard University and Brigham and Women’s Hospital from the National Center for Research Resources. Benjamin E. Gewurz was supported by a Broad Institute Founder’s grant. Yifei Liao was supported by a Lymphoma Research Foundation award. Carl D. Langefeld was supported by P01 AR49084, NIAMS, Program Project in the Genetics of SLE (UAB-SLE2), and P01 AI083194, NIH, Genomics of Lupus (SLEGEN P01). This work was supported by the Cell Discovery Network, a collaborative initiative funded by the Manton Foundation and the Warren Alpert Foundation at Boston Children’s Hospital.

## Supplementary material

### Figures

**Figure S1:**
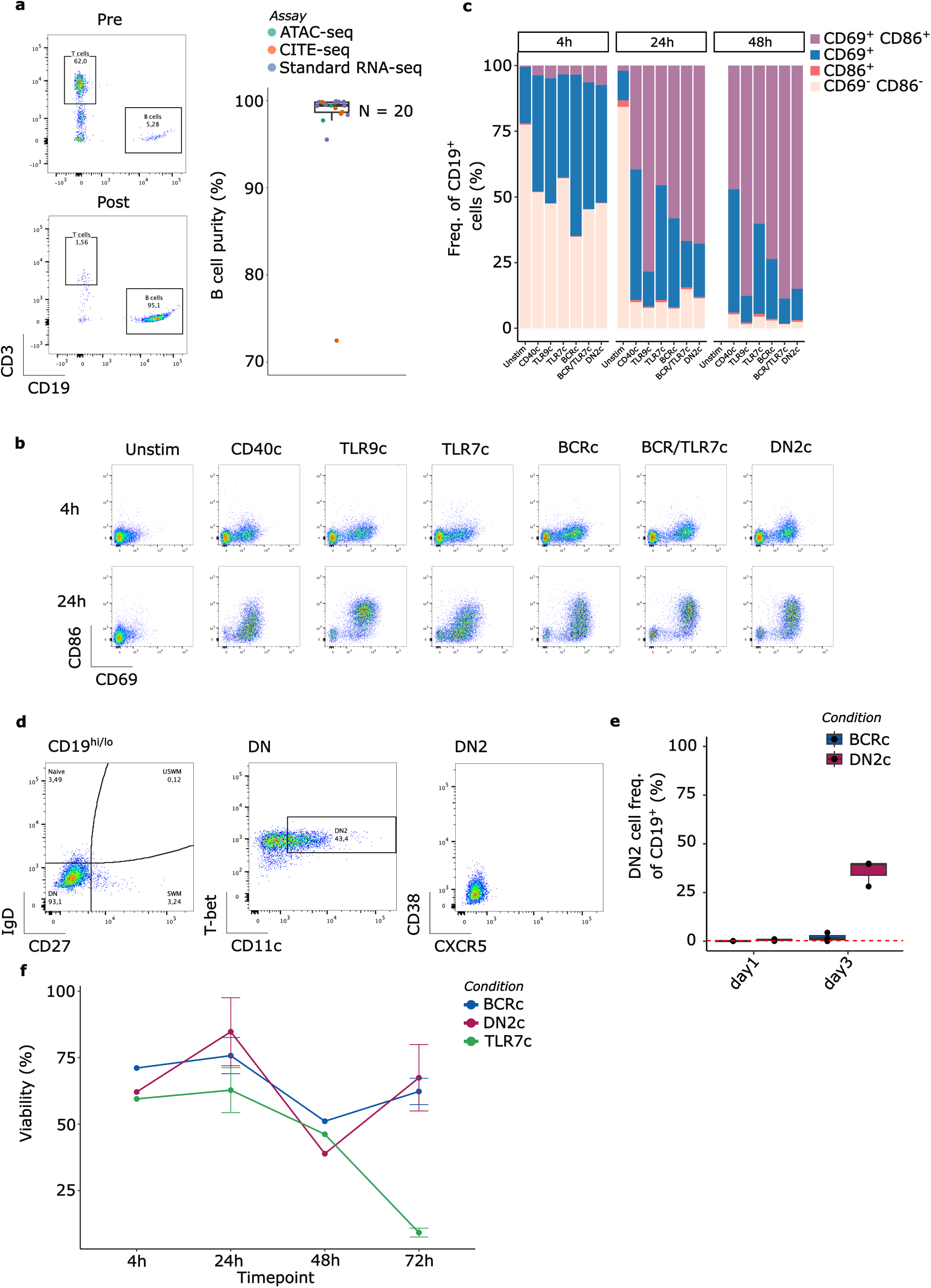
**a**, Surface staining of B cells for the indicated markers before (top) and after (bottom) enrichment. On the right proportion of B cells after enrichment across assays (n = 20). **b**, Expression of activation markers CD69 and CD86 on B cells after 4 hours and 24 hours of culture under the indicated conditions. **c**, Frequency of active and non-active cells across conditions and timepoints. **d**, Gating strategy for identifying DN2 B cells based on surface marker expression at 72 hours post-culture. **e**, Frequency of DN2 B cells at 72 hours after culture (n = 3). **f**, Viability of B cells over time under the indicated conditions.

**Figure S2:**
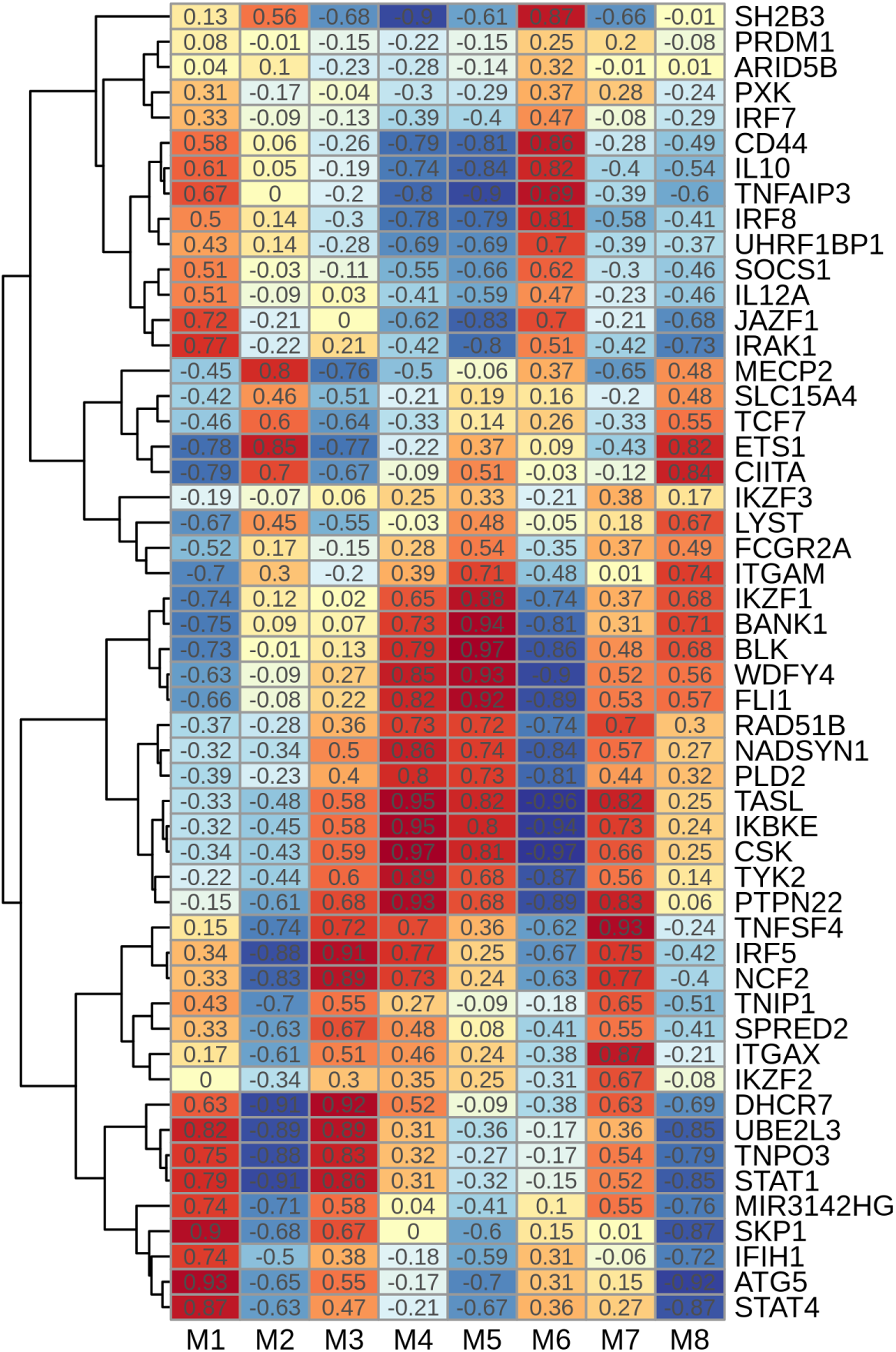
Module membership of SLE candidate genes. Colors represent the WGCNA kME value.

**Figure S3:**
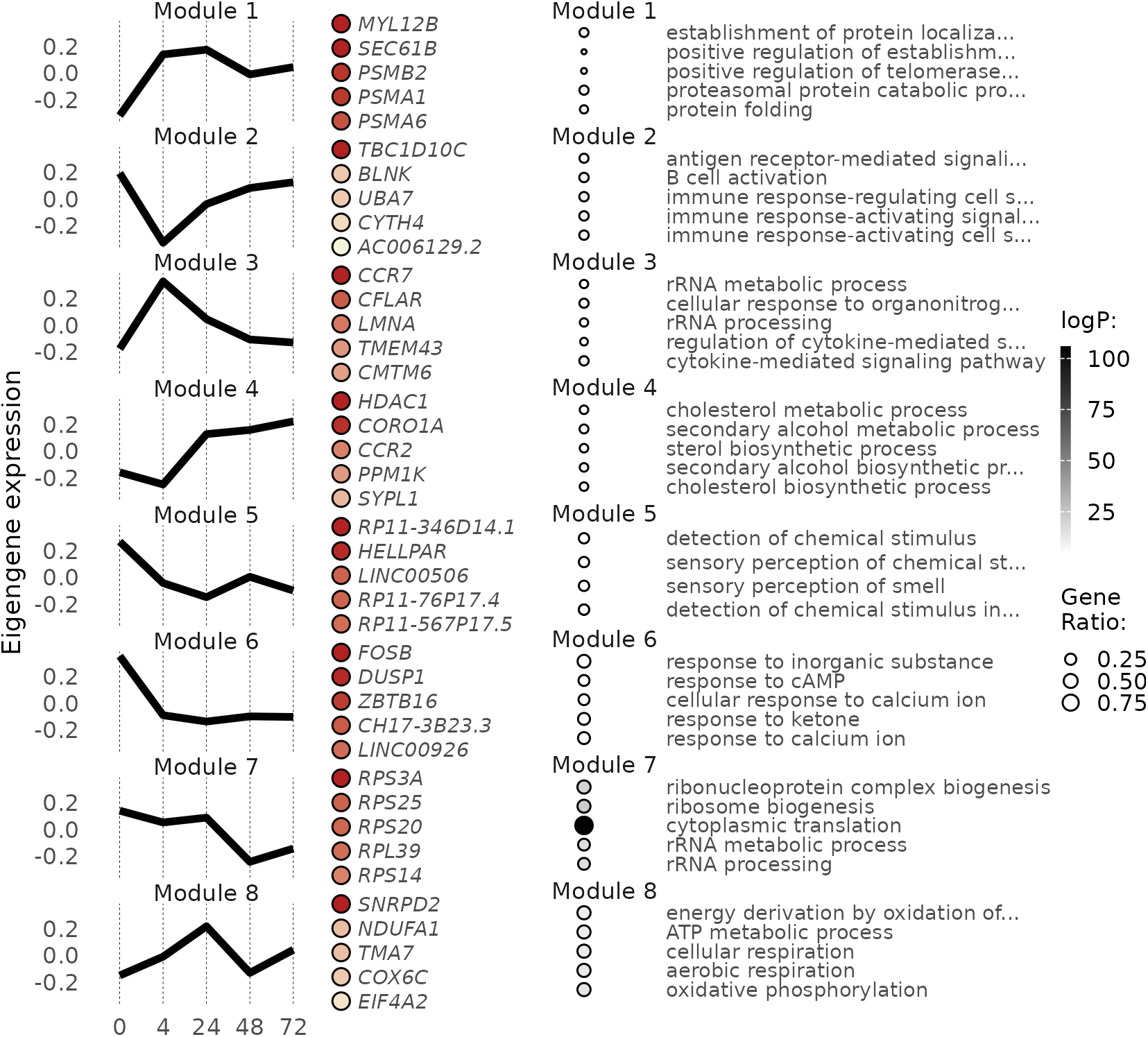
WGCNA modules (left) and gene ontology enrichments (right) in the CD40c condition.

**Figure S4:**
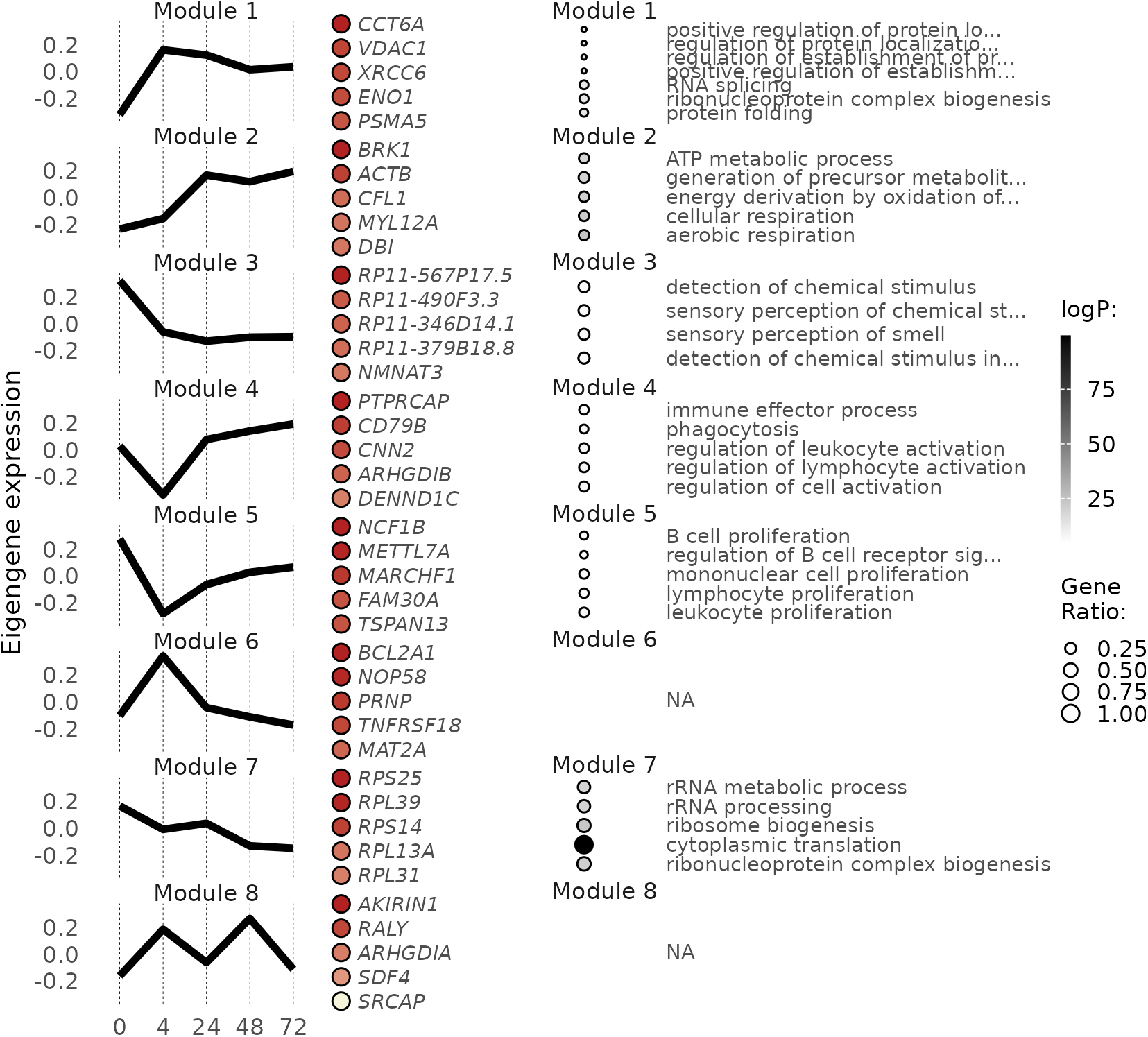
WGCNA modules (left) and gene ontology enrichments (right) in the TLR7c condition.

**Figure S5:**
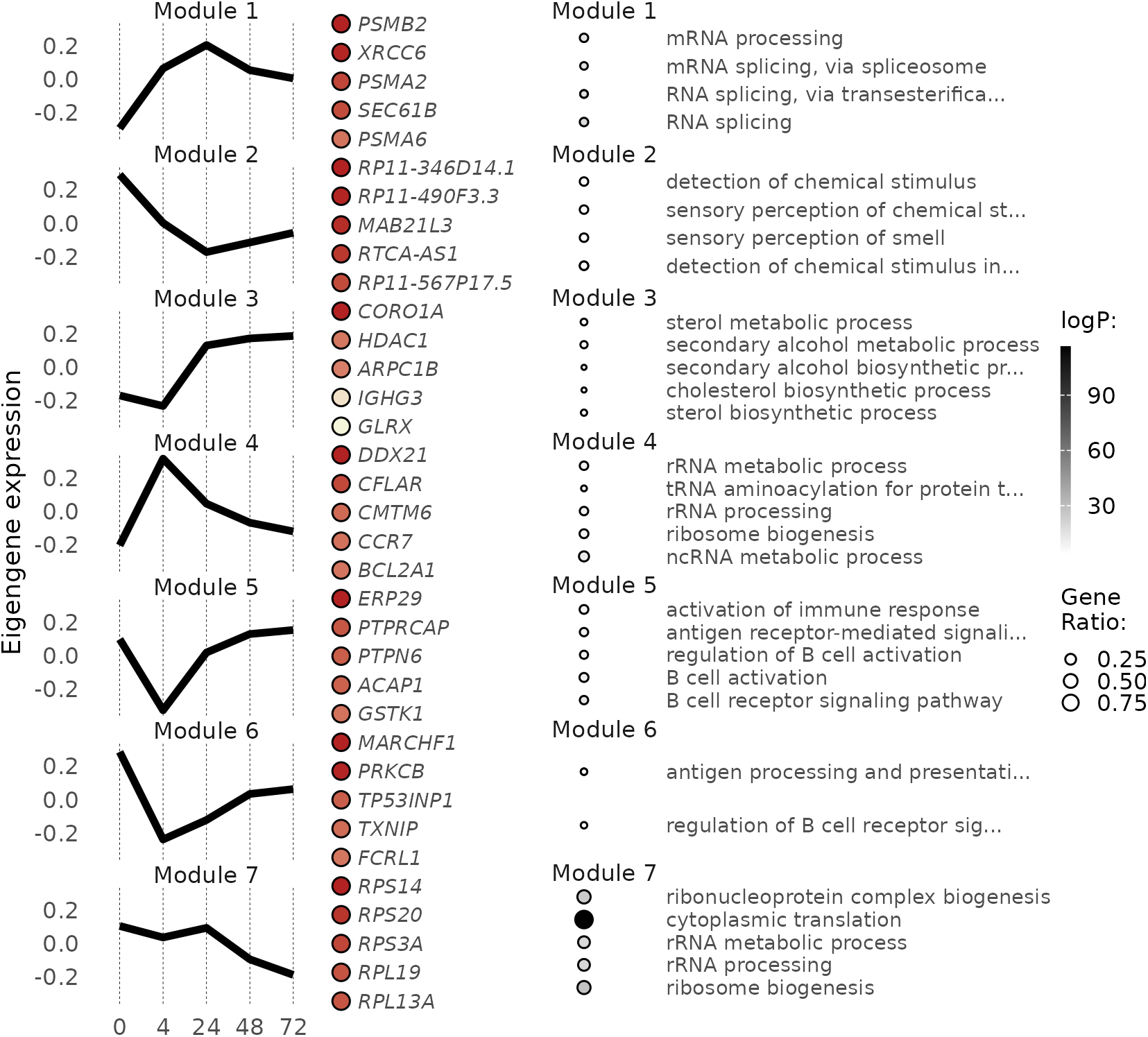
WGCNA modules (left) and gene ontology enrichments (right) in the TLR9c condition.

**Figure S6:**
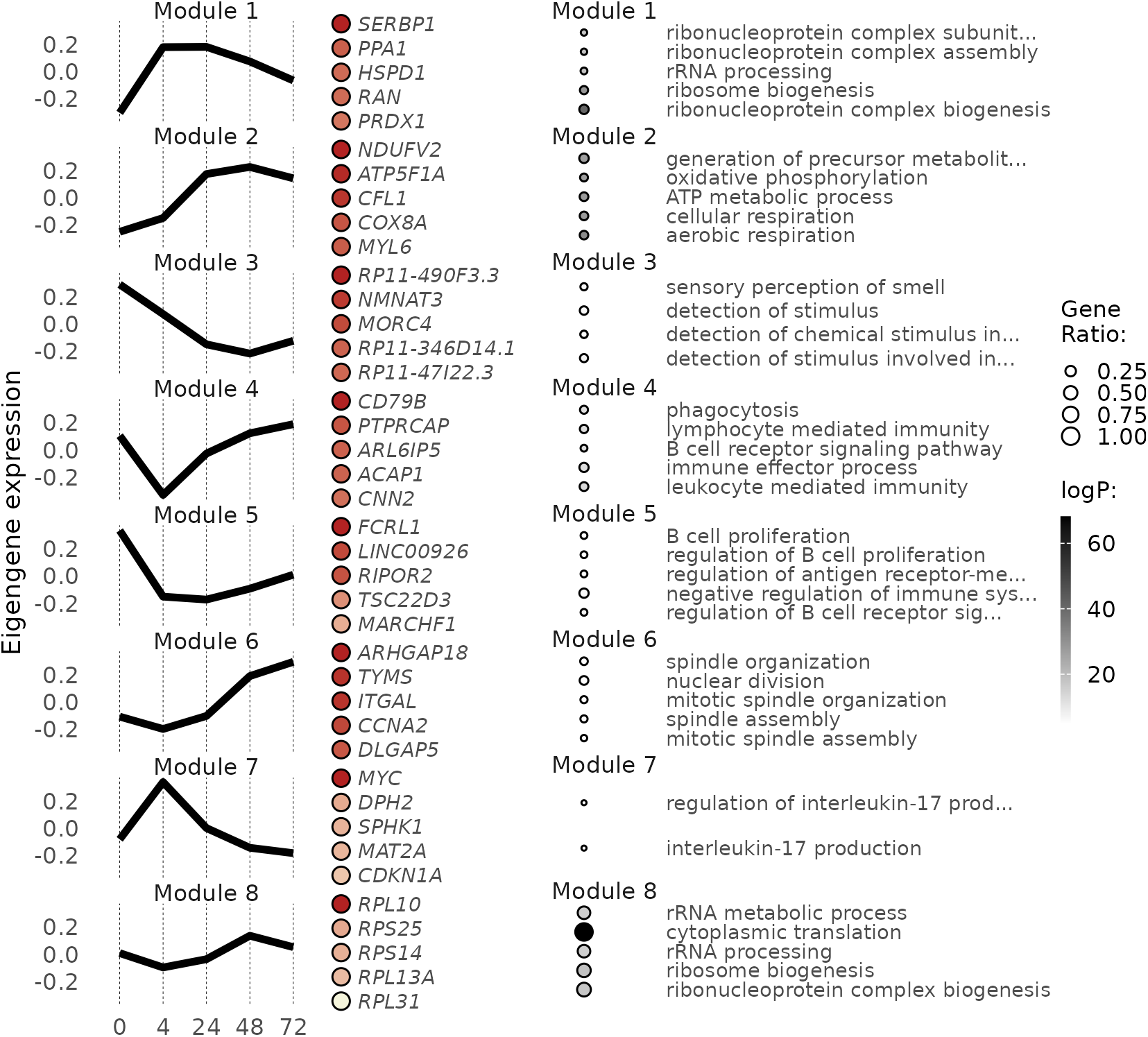
WGCNA modules (left) and gene ontology enrichments (right) in the BCRc condition.

**Figure S7:**
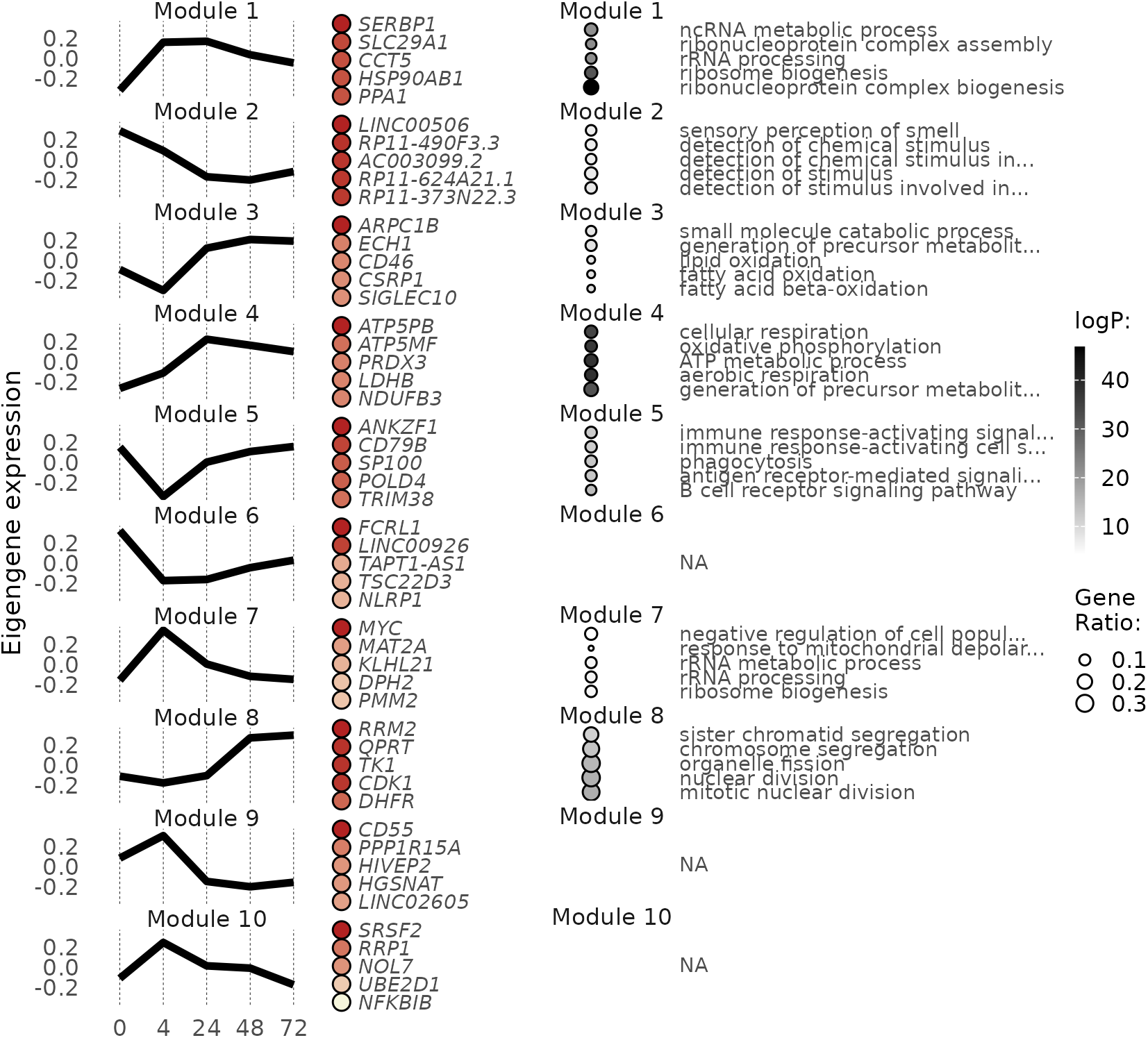
WGCNA modules (left) and gene ontology enrichments (right) in the BCR/TLR7c condition.

### Table legends

**Table S1:** Gene Ontology enrichment analysis for genes overexpressed in DN2c with respect to BCRc at 72 hours at 1% FDR.

## Notes

### Competing Interest Statement

The authors have declared no competing interest.

### Author Declarations

Institutional Review Boards of Mass General Brigham and Boston Childrens Hospital gave ethical approval for this work.

### Summary of Updates

No changes in the original Introduction, Results and Methods sections were implemented. In this revision, we put main figures in place, we added a link to the abstract to facilitate data access, and improve Discussion.

